# C-reactive protein for pulmonary tuberculosis screening and treatment response monitoring in children

**DOI:** 10.1101/2025.04.24.25326338

**Authors:** Joy Githua, Jerphason Mecha, Joshua Stern, Jaclyn N. Escudero, Lilian Njagi, Lucy Kijaro, Jacqueline Mirera, Wilfred Murithi, Grace John-Stewart, Elizabeth Maleche-Obimbo, Videlis Nduba, Sylvia M. LaCourse

## Abstract

C-reactive protein (CRP) was investigated as a marker for diagnostic screening and monitoring response to treatment for tuberculosis (TB) in Kenyan children. For screening, CRP levels did not differ significantly between children with versus without TB. However, median CRP levels decreased significantly during TB treatment in confirmed (p=0.03) and unconfirmed TB (p=0.002) suggesting potential of treatment response monitoring.

**SUMMARY:** CRP levels did not differ significantly between children with versus without TB at diagnosis but decreased significantly during TB treatment in confirmed (p=0.02) and unconfirmed TB (p<0.001). While CRP’s diagnostic screening performance was suboptimal, findings suggest potential utility for treatment response monitoring in children with TB.

## INTRODUCTION

Most TB-related pediatric deaths occur in children under five, primarily due to missed diagnoses.^1^ Young children frequently present with disseminated or extrapulmonary TB, making diagnosis challenging through conventional respiratory sampling methods.^2^ Limitations of current respiratory-based diagnostic methods, including culture and polymerase chain reaction (PCR)-based tests, highlight the necessity for non-sputum-based diagnostics for pediatric TB detection and treatment response monitoring.^3^

C-reactive protein (CRP), an acute-phase protein associated with infectious and inflammatory conditions, including TB, has shown potential for screening and monitoring treatment response, mainly in adult TB. ^4^ However, its utility in pediatric TB management remains under-researched.^5^ We evaluated CRP as a diagnostic screening and treatment response monitoring tool in a cohort of Kenyan children presenting with symptoms suggestive of TB.

## METHODS

Our prospective observational study enrolled children ≤15 years presenting to the National Tuberculosis Program and HIV care clinics in Nairobi, Kenya, with suggestive TB symptoms (including persistent cough, fever, night sweats, weight loss / poor weight gain, lethargy) . We focused on children highly suspected of having TB or recommended for treatment. Children treated for TB for more than seven days or planning to leave Nairobi County were excluded.

Participants underwent comprehensive evaluation including interviews, medical record review, physical examination, HIV testing, chest radiographs (CXRs), and respiratory specimens (two samples by induced or expectorated sputum or gastric aspirates for children <5 years) for testing by Xpert Ultra, microscopy, and culture. CXRs were interpreted by a radiologist and study clinician with an additional review by a pediatric pulmonologist in case of discrepancies. They were classified as suggestive or not suggestive of TB and categorized based on pathology using standardized forms. TB treatment followed national guidelines, which were 6 months of treatment for drug-sensitive TB at the time of this study. Follow-up visits were scheduled at 2 weeks and months 1, 2, 4, 6, 12, and 24. Participants were categorized using NIH consensus definitions as confirmed TB (positive Xpert/Ultra and/or culture), unconfirmed TB (symptoms consistent with TB plus supporting evidence of TB but negative Xpert and culture), or unlikely TB (symptoms present but not meeting unconfirmed TB criteria).^6^

We measured CRP levels at baseline and during follow-up visits for participants on TB treatment. CRP was considered positive if >5 mg/L, with a lower detection limit of 0.6 mg/L. Due to reporting error a subset of samples (57 samples from 54 participants) during 1 month of testing with levels of 2.5 mg/L or below were erroneously reported by the laboratory as 2.5 mg/L. This did not affect the diagnostic performance analysis using the 5 mg/L cut-off but could potentially impact quantitative CRP analyses. To address this, we performed sensitivity analyses both excluding the 57 incorrectly reported samples and additionally assigning these values a midpoint value of 1.25 mg/L (0-2.5 mg/L) for quantitative analyses. ‘Near treatment end’ was defined as completing at least 4 months of TB treatment.

Demographic and clinical characteristics were summarized. CRP performance for TB diagnosis was evaluated using 5 mg/L and 10 mg/L cut-offs.^7^ Correlates of CRP positivity (≥5 mg/L) were assessed using a generalized linear model (GLM) with log link and Poisson family. The final model included variables with p<0.10 in univariate analysis. Prevalence ratios (PRs) were calculated using chi-square and Wilcoxon rank sum tests. Serial CRP was compared between baseline and near treatment end for children who had received ≥4 months of TB treatment using McNemar’s chi-square and signed rank tests.

We further estimated CRP’s accuracy for TB diagnosis by calculating area under the curve (AUC) for receiver operating characteristics (ROC) comparing 1) children with confirmed TB to children with unlikely TB; 2) confirmed TB (microbiologic reference standard, MRS) to unconfirmed and unlikely TB; and 3) confirmed and unconfirmed TB (composite reference standard, CRS) to unlikely TB.

### Ethics

This study was approved by the University of Nairobi/Kenyatta National Hospital Ethics and Research Committee, Kenya Medical Research Institute Scientific Ethics Research Unit, and University of Washington Institutional Review Board. We obtained parental consent from all enrolled children and assent from those aged >13 years.

## RESULTS

Between May 2022 to July 2023, 467 children were screened for eligibility, and 330 participants were enrolled **(Supplemental Figure 1)**. Of these, 292 (88.5%) children with CRP results were included in the analysis. The 292 children had a median age 3.0 years (interquartile range (IQR): 1.0-5.0 years), 46.2% were male (n=135), and 3.1% (n=9) were living with HIV **(Supplemental Table 1)**. Eighteen (6.1%) had confirmed TB, 222 (76.0%) unconfirmed TB, and 52 (17.8%) unlikely TB.

Baseline CRP levels in confirmed (median 3.8 mg/L, p=0.18), unconfirmed (median 1.8 mg/L, p=0.92), showed no significant differences when comparing each group versus unlikely TB (median 1.6 mg/L) **(Table 1)**. Sensitivity analysis excluding 57 participants with erroneously reported CRP values (≤2.5 mg/L) and setting them to 1.25 mg/L yielded similar results **(Supplemental Table 2)**.

**Table 1:** Screening Performance of CRP in Pulmonary TB and Treatment Response Monitoring (pre vs. near end treatment) for Children.

| **Diagnostic screening** |  |  |  |
| --- | --- | --- | --- |
| **N=292** | **Confirmed TB<sup>a</sup>**<br>**n=18** | **Unconfirmed TB<sup>a</sup>**<br>**n=222** | **Unlikely TB<sup>a</sup>**<br>**n=52** |
| CRP ≥5 mg/L | 9 (50.0 %) | 77 (34.7 %) | 15 (28.8 %) |
| P-Value* | 0.10 | 0.42 | Reference |
| CRP ≥10 mg/L | 8 (44.4 %) | 60 (27.0%) | 14 (26.9 %) |
| P-Value* | 0.17 | 0.99 | Reference |
| CRP (Median, IQR) | 3.8 (0.5, 47.8) | 1.8 (0.5, 12.5) | 1.6 (0.5, 11.6) |
| P-Value* | 0.18 | 0.92 | Reference |
| **Treatment response\*\*** |  |  |  |
| **N=95** | **Confirmed TB**<br>**n=11** | **Unconfirmed TB**<br>**n=79** | **Unlikely TB**<br>**n=5** |
| Pre TBtx-CRP mg/L<br>(Median, IQR) | 8.1 (0.4, 54.0) | 2.4 (0.7, 13.5) | 2.0 (1.1, 4.5) |
| Near TBtx end CRP<br>mg/L (Median, IQR) | 0.5 (0.2, 1.2) | 0.7 (0.3, 2.5) | 1.6 (0.7, 2.5) |
| P-Value*** | **0.02** | **<0.001** | 0.22 |
| **Abbreviations:** IQR, Interquartile range<br><sup>a</sup> Based on international consensus clinical case definitions for pediatric TB via post-hoc classification.<br>* Confirmed or Unconfirmed TB compared to Unlikely TB<br>** Among participants who completed at least 4 months of TB treatment<br>*** Pre TB treatment compared to near treatment end<br>Median values were compared using Wilcoxon signed rank-sum test<br>Sensitivity/specificity comparisons were done using Mc-Nemar's chi square test |  |  |  |

At 5 mg/L threshold, CRP sensitivity was 50.0% (95%CI 26.0-74.0) for confirmed and 34.7% (95%CI 28.4-41.3) for unconfirmed TB, and specificity was 71.2% (95%CI 58.7-71.6) for unlikely TB **(Table 1)**. At 10 mg/L threshold, sensitivity decreased to 44.4% (95%CI 21.5-69.2) for confirmed and 27.0% (95%CI 21.3-33.4) for unconfirmed TB, and specificity was 73.1% (95%CI 66.6-78.7) among children with unlikely TB.

Among 95 children who completed at least 4 months of TB treatment with at least two serial CRP measures, the median CRP was higher pre-treatment compared to near treatment end among children with confirmed (8.1 vs. 0.5 mg/L; p=0.02) and unconfirmed TB (2.4 vs. 0.7 mg/L; p < 0.001), but not unlikely TB (2.0 vs. 1.6 mg/L; p=0.35) **(Table 1)**, with sensitivity analysis 1) setting them to 1.25 mg/L 2) and excluding the erroneous values, yielding similar results **(Supplemental Table 2)**.

Baseline positive QFT was associated with a 1.5 times higher risk of baseline CRP positivity. CLHIV had a doubled risk of CRP positivity. CXR suggestive of TB, confirmed TB, and TB treatment initiation, were associated with increased CRP positivity risk in univariate analysis but these associations were not statistically significant after adjustment (**Supplemental Table 3)**.

We performed ROC analysis for CRP performance using different reference standards (**Supplemental Table 4)**. For confirmed TB vs. unlikely TB patients, AUC was 0.61 (95% CI: 0.45-0.77) with a cut-off value of 2.28 mg/L. For the microbiologic reference standard (MRS) (confirmed vs. unconfirmed + unlikely TB), AUC was 0.59 (95% CI: 0.45-0.74) with a cut-off value of 2.44 mg/L. For the composite reference standard (CRS, confirmed + unconfirmed vs. unlikely TB), AUC was 0.51 (95% CI: 0.43-0.60) with a 2.10 mg/L cut-off value.

## Discussion

CRP has emerged as a potential biomarker for active TB and treatment response monitoring primarily in adults. Although screening diagnostic performance was suboptimal in children in our study, CRP levels significantly decreased between TB treatment initiation and near completion in those with confirmed and unconfirmed TB, suggesting potential utility for treatment response. Among adults with TB, results have generally met the WHO criteria for a screening test with ≥90% sensitivity and ≥70% specificity.^4,5^ However, our study’s sensitivity did not meet WHO criteria at the 5 mg/L or 10 mg/L thresholds, consistent with prior pediatric studies^4,5^ highlighting a performance divergence between children and adults.

Our treatment response results are consistent with previous findings. In a systematic review conducted by Zimmer et al. in individuals >15 years of age, a significant (76.1%) reduction in CRP 8 weeks post-treatment initiation, with further decline, was observed.^8^ Similarly, Wilson et al. reported a significant decrease in CRP levels among adults with presumptive TB, in the confirmed and probable TB groups compared to the unlikely TB group.^9^ Additionally, our results concur with previous research showing a correlation between high CRP levels and HIV positivity,^4,10^ and radiological severity of the disease.^10^

Similar to a study in Uganda^5^ our ROC analysis revealed moderate diagnostic performance of CRP in distinguishing between confirmed TB and unlikely TB cases in children, with an AUC of 0.61 and MRS demonstrated moderate diagnostic performance with an AUC of 0.59. However, using CRS resulted in an AUC of 0.51 revealing close to random chance for TB diagnosis.

Strengths of this study include comprehensive investigations, standardized longitudinal follow-up, adherence to consensus guidelines for TB classification, and standardized CRP cut-offs. Limitations include inaccurate CRP reporting for 57 participants, which could have resulted in bias. However, sensitivity analyses excluding these assays and using 1.25mg/L as a mid-point value showed no significant differences in findings. The high predominance of unconfirmed TB cases suggests some treated individuals may have had alternative conditions. Despite this, thorough evaluations were conducted following TB classification guidelines.

Another limitation was the exclusion of 38 participants (11.5%) due to missing CRP results. However, these participants were similar in key characteristics (e.g., age, gender, HIV status, nutritional status, and TB symptoms) to those with CRP results, minimizing bias in our findings.

## Conclusion

CRP fell short of WHO triage test targets for TB but showed promise for monitoring treatment response in children, necessitating further evaluation. Combined with other pediatric TB algorithms, CRP may be valuable for diagnosing TB in children, where TB is often paucibacillary, and for monitoring treatment response.

## Supporting information

Supplemental Figure 1_ Participant Flow Chart

Supplemental Table 1: Baseline Characteristics

Supplemental Table 2_ Sensitivity Analysis

Supplemental Table 3: Correlates of CRP Positivity

Supplemental Figure 2: ROC for CRP to diagnose TB

Supplemental Table 4: AUC under the ROC curve for CRP levels

## Data Availability

All data supporting the findings of this study are fully available within the manuscript and its supplementary materials

## AUTHOR CONTRIBUTIONS

S.M.L. designed the parent study. S.M.L.,V.N., and E.M.O. developed the parent study protocol. S.M.L. is the principal investigator and V.N is the site principal investigator. S.M.L., J.S. J.N.E were responsible for the statistical design of the study. J.S., J.M., J.G., and S. M. L. performed the data analysis and developed the tables and figures. J.G., S.M.L., and V.N. prepared the initial draft of the manuscript. All authors have read and approved the manuscript.

## ACKNOWLEDGEMENTS

The authors acknowledge the KEMRI-CRDR study staff at the University of Nairobi/Kenyatta National Hospital, Nairobi County TB clinics, and the referral hospitals.

Above all, we offer sincere thanks to the study participants.

## Funding

This work was supported by the NIH/National Institute of Allergy and Infectious Diseases R01AI162152 and R01AI179714, S.M.L., NIH grant P30 AI027757; University of Washington/Fred Hutch, Center for AIDS Research (CFAR) New Investigator Award (NIA) CFAR-NIA, J.G., (AI027757); TBHTP (Tuberculosis and HIV Co-infection Training Program)-D43-TW011817, J.G.

## Conflicts of Interest

The authors declare no conflicts of interest

## Data Access Statement

**Supplemental Digital Content 1: Figure 1: Participant flowchart**

**Supplemental Digital Content 2: Supplemental Table 1 : Baseline characteristics**

**Supplemental Digital Content 3: Supplemental Table 2 : Sensitivity Analysis Diagnosis and Treatment Response**

**Supplemental Digital Content 4: Supplemental Table 3 : Correlates of CRP positivity at baseline**

**Supplemental Digital Content 5: Supplemental Figure 2: Receiver operating characteristic (ROC) for C-reactive protein to diagnose TB**

**Supplemental Digital Content 6: Supplemental Table 4: Area under the ROC Curve for C-Reactive Protein Levels toDetect Childhood TB**

## Notes

### Competing Interest Statement

The authors have declared no competing interest.

### Author Declarations

Scientific Ethics Review Unit of Kenya Medical Research Institute gave ethical approval for this work University of Washington IRB gave ethical approval for this work University of Washington IRB gave ethical approval for this work Kenyatta National Hospital-University of Nairobi Ethics Review Committee gave ethical approval for this work

