## Supplementary figures and images for "C-reactive protein for pulmonary tuberculosis screening and treatment response monitoring in children"

### Supplemental Figure 1_ Participant Flow Chart

**Supplemental Figure 1: Par****ticipant flowchart**

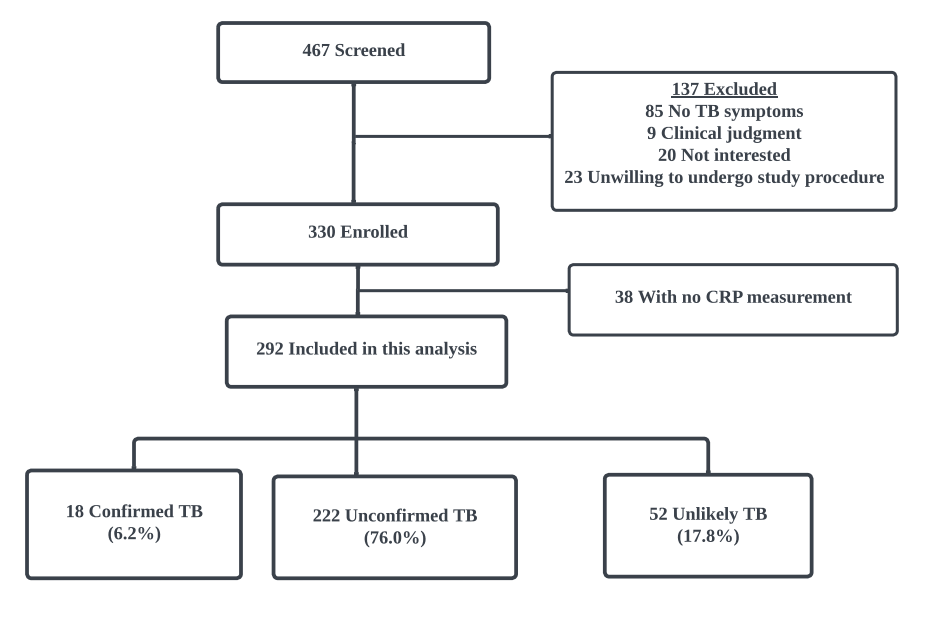
