## Supplemental Table 1: Baseline Characteristics for "C-reactive protein for pulmonary tuberculosis screening and treatment response monitoring in children"

| **Supplemental Table 1 : Baseline characteristics** | | | | |
| --- | --- | --- | --- | --- |
|  | **Overall** | **Confirmed TB ^a^** | **Unconfirmed**  **TB ^a^** | **Unlikely**  **TB ^a^** |
|  | **N=292** | **N=18** | **N=222** | **N=52** |
|  | **Median (IQR) or n (%)** | **Median (IQR) or n (%)** | **Median (IQR) or n (%)** | **Median (IQR) or n (%)** |
| **Demographics** | | | | |
| Age (years) | 3.0 (1.0, 5.0) | 1.5 (1.0, 8.0) | 2.0 (1.0, 5.0) | 4 (1.5, 8.5) |
| Female sex | 157 (53.8) | 11 (61.1) | 116 (52.3) | 30 (57.7) |
| **Clinical Presentation** | | | | |
| BMIz (N=289) | -0.4 (-1.8, 0.5) | -0.7 (-1.2, -0.0) | -0.3 (-1.8, 0.5) | -0.5 (-2.1, 0.5) |
| Underweight^b^  (BMIz<-2) (N=289) | 64 (22.1) | 2 (11.1) | 48 (21.8) | 14 (27.5) |
| WHZ (N=203)**^c^** | -0.7 (-2.1, 0.3) | -0.8 (-1.4, -0.4) | -0.5 (-2.1, 0.4) | -1.4 (-2.8, 0.8) |
| Wasted (WHZ<-2) (N=203)**^d^** | 58 (28.6) | 2 (16.7) | 42 (26.2) | 14 (45.2) |
| WAZ (N=204)^c^ | -1.4 (-2.4, -0.4) | -1.9 (-2.6, -0.6) | -1.2 (-2.3, -0.3) | -1.9 (-2.9, -0.4) |
| Underweight  (WAZ<-2) (N=204)**^d^** | 73 (35.8) | 5 (41.7) | 53 (32.9) | 15 (48.4) |
| CLHIV | 9 (3.1) | 0 (0.0) | 8 (3.6) | 1 (1.9) |
| **TB Features** | | | | |
| NIH criteria signs/symptoms of TB **^e^** | 238 (81.5) | 16 (88.9) | 190 (85.6) | 32 (61.5) |
| TST positive (N=288) | 104 (36.1) | 10 (55.6) | 83 (37.9) | 11 (21.6) |
| QFT Positive (N=222) | 36 (12.3) | 5 (27.8) | 27 (12.2) | 4 (7.7) |
| TB contact | 117 (40.1) | 10 (55.6) | 98 (44.1) | 9 (17.3) |
| CXR suggestive of TB | 185 (63.4) | 16 (88.9) | 166 (74.8) | 3 (5.8) |
| Culture positive(N=261) **^f^** | 9 (3.4) | 9 (52.9) | -- | -- |
| Xpert positive(N=276) **^f^** | 15 (5.4) | 15 (83.3) | -- | -- |
| Urine LAM positive (N=166) | 17 (10.2) | 1 (10.0) | 14 (11.1) | 2 (6.7) |
| TB treatment-initiated **^g^** | 127 (43.5) | 15 (83.3) | 103 (46.4) | 9 (17.3) |
| Positive response TB tx (N=127) **^h^** | 116 (91.3) | 13 (86.7) | 94 (91.3) | 9 (100.0) |
| **Abbreviations**: IQR: Interquartile range; WAZ ,weight for-age z score; WHZ, weight for-height z score; BMI, Body Mass Index (BMI) –for-age z score ; HUU, HIV unexposed uninfected; HEU, HIV exposed uninfected; CLHIV, children living with HIV; NIH, National Institutes of Health; TST, tuberculin skin test; QFT, QuantiFERON test; CXR, chest radiograph; Mtb, mycobacterium tuberculosis; Xpert, Xpert MTB/ULTRA; LAM, lipoarabinomannan; TBTx, TB treatment.  **N:** Number of participants with results; **n:** number of participants with positive results  ^a^ Based on international consensus clinical case definitions for pediatric TB via post-hoc classification.  ^b^ BMI z<-2 For the entire population  ^c^ Among children 5 years and under  ^d^ Among children 5 years and under: WHZ <-2 or MUAC <12·5 cm or WAZ<-2 r  ^e^ Persistent cough (>14 days), fever (>7 days), failure to thrive, or lethargy (>7 days)· Failure to thrive=wasted (WHZ<-2 or MUAC<12·5) or underweight (WHZ<-2) or Thinness (BMI <-2) at enrollment (growth trajectories unavailable before enrollment). ^f^ Sputum or gastric aspirate.  ^g^ Received TB treatment at enrollment or 2-weeks after enrollment  ^h^ Positive response to TB treatment after 2 weeks of enrollment | | | | |
