## Supplemental Table 2_ Sensitivity Analysis for "C-reactive protein for pulmonary tuberculosis screening and treatment response monitoring in children"

**Supplemental Table 2: Sensitivity Analysis of Screening Performance of CRP in Pulmonary TB and Treatment Response Monitoring (pre vs. near end treatment) for Children**

| **Diagnostic screening: Setting to 1.25mg/L^¥^** | | | |
| --- | --- | --- | --- |
| **N=292** | **Confirmed TB^a^**  **n=18** | **Unconfirmed TB^a^ n=222** | **Unlikely TB^a^**  **n=52** |
| CRP ≥5 mg/L | 9 (50.0 %) | 77 (34.7 %) | 15 (28.8 %) |
| P-Value* | 0.10 | 0.42 | Reference |
| CRP ≥10 mg/L | 8 (44.4 %) | 60 (27.0%) | 14 (26.9 %) |
| P-Value* | 0.17 | 0.99 | Reference |
| CRP (Median, IQR) | 3.8 (0.5, 47.8) | 1.3 (0.5, 12.5) | 1.3 (0.5,11.6) |
| P-Value* | 0.19 | 0.94 | Reference |
| **Diagnostic screening: Exclusion of 2.5mg/L^#^** | | | |
| **N=279** | **Confirmed TB^a^**  **n=16** | **Unconfirmed TB^a^ n=214** | **Unlikely TB^a^**  **n=49** |
| CRP ≥5 mg/L | 8 (50.0 %) | 77 (36.0 %) | 15 (30.6 %) |
| P-Value* | 0.16 | 0.48 | Reference |
| CRP ≥10 mg/L | 7 (43.8 %) | 60 (28.0%) | 14 (28.6 %) |
| P-Value* | 0.26 | 0.94 | Reference |
| CRP (Median, IQR) | 3.8 (0.4, 50.9) | 1.6 (0.5, 12.9) | 1.4 (0.5, 12.4) |
| P-Value* | 0.23 | 0.93 | Reference |
| **Treatment Response**: Setting to 1.25mg/L^¥^** | | | |
| **N=95** | **Confirmed TB^a^**  **n=11** | **Unconfirmed TB^a^ n=79** | **Unlikely TB^a^**  **n=5** |
| Pre TBtx-CRP mg/L (Median, IQR) @ 1.25 | 8.1 (0.4, 54.0) | 1.5 (0.7, 12.8) | 1.4 (1.3, 4.5) |
| Near TBtx end CRP mg/L (Median, IQR) | 0.5 (0.2, 1.3) | 0.8 (0.4, 2.5) | 0.8 (0.7, 2.4) |
| P-Value*** | **0.02** | **0.0001** | 0.35 |
| **Treatment response**: Exclusion of 2.5mg/L^#^** | | | |
| **N=77** | **Confirmed TB^a^**  **n=10** | **Unconfirmed TB^a^ n=62** | **Unlikely TB^a^**  **n=3** |
| Pre TBtx-CRP mg/L (Median, IQR) | 10.0 (0.4, 54.0) | 1.8 (0.7, 14.3) | 4.5 (1.4, 18.6) |
| Near TBtx end CRP mg/L (Median, IQR) | 0.6 (0.2, 1.3) | 0.7 (0.4, 1.4) | 0.8 (0.7, 2.4) |
| P-Value*** | **0.04** | **0.0001** | 0.29 |
| **Abbreviations**: IQR, Interquartile range  ^a^ Based on international consensus clinical case definitions for pediatric TB via post-hoc classification.  * Confirmed or Unconfirmed TB compared to Unlikely TB  ** Among participants who completed at least 4 months of TB treatment  *** Pre TB treatment compared to near treatment end  ¥- Setting CRP levels inadvertently reported as 2.5mg/L to 1.25mg/L  # Excluding participants whose result was inadvertently reported as 2.5mg/L for levels of 2.5 mg/L  Median values were compared using Wilcoxon signed rank-sum test  Sensitivity/specificity comparisons were done using Mc-Nemar’s chi square test | | | |
