## Supplemental Table 3: Correlates of CRP Positivity for "C-reactive protein for pulmonary tuberculosis screening and treatment response monitoring in children"

| **Supplemental Table 3: Correlates of CRP positivity at baseline (CRP ≥5 mg/L)** | | | | | | | |
| --- | --- | --- | --- | --- | --- | --- | --- |
| **Baseline characteristics** | **Overall** | **CRP positive**  **N=101**  **n (%) or median (IQR)** | **CRP negative**  **N=191**  **n (%) or median (IQR)** | **RR (95% CI)^*^** | **p** | **aRR (95% CI)^Ϊ^** | **p** |
|  | **N=292** |  |  |  |  |  |  |
|  | **n (%) or median (IQR)** |  |  |  |  |  |  |
| **Demographics** | | | | | | | |
| Age (years) | 3.0 (1.0, 5.0) | 3.0 (1.0, 5.0) | 2.0 (1.0, 5.0) | 1.01 (0.97-1.05) | 0.636 |  |  |
| Female sex | 157 (53.8) | 59 (58.4) | 98 (51.3) | 1.21 (0.87-1.67) | 0.251 |  |  |
| **Clinical Presentation** | | | | | | | |
| BMIz (N=289) | -0.4 (-1.8, 0.5) | -0.7 (-2.0, 0.3) | -0.3 (-1.7, 0.5) | 0.94 (0.87-1.02) | 0.114 |  |  |
| Underweight^a^  (BMIz<-2) (N=289) | 64 (22.1) | 26 (25.7) | 38 (20.2) | 1.22 (0.86-1.73) | 0.268 |  |  |
| WHZ (N=203)^b^ | -0.7 (-2.1, 0.3) | -0.8 (-2.3, 0.3) | -0.5 (-2.1, 0.4) | 0.97 (0.88-1.08) | 0.560 |  |  |
| Wasted (WHZ<-2) (N=203)^c^ | 58 (28.6) | 21 (30.4) | 37 (27.6) | 1.09 (0.72-1.65) | 0.671 |  |  |
| WAZ (N=204) ^b^ | -1.4 (-2.4, -0.4) | -1.7 -2.6, -0.4) | -1.2 (-2.3, -0.4) | 0.94 (0.83-1.07) | 0.382 |  |  |
| Underweight  (WAZ<-2)(N=204)^c^ | 73 (35.8) | 29 (42.0) | 44 (32.6) | 1.30 (0.89-1.91) | 0.179 |  |  |
| HIV status |  |  |  |  |  |  |  |
| HUU | 275 (94.2) | 92 (91.1) | 183 (95.8) | Reference |  | Reference |  |
| HEU | 8 (2.7) | 3 (3.0) | 5 (2.6) | 1.21 (0.45-2.79) | 0.806 | 1.27 (0.56-2.88) | 0.563 |
| CLHIV | 9 (3.1) | 6 (5.9) | 3 (1.6) | 1.88 (1.22-3.26) | **0.006** | 1.93 (1.17-3.19) | **0.010** |
| **TB Features** | | | | | | | |
| NIH criteria signs/symptoms of TB^d^ | 238 (81.5) | 80 (79.2) | 158 (82.7) | 0.86 (0.59-1.26) | 0.452 |  |  |
| TB Classification |  |  |  |  |  |  |  |
| Confirmed TB | 18 (6.2) | 9 (8.9) | 9 (4.7) | 1.73 (0.92-3.25) | **0.087** | 1.14 (0.56-2.33) | 0.719 |
| Unconfirmed TB | 222 (76.0) | 77 (76.2) | 145 (75.9) | 1.20 (0.76-1.91) | 0.436 | 0.89 (0.50-1.58) | 0.696 |
| Unlikely TB | 52 (17.8) | 15 (14.9) | 37 (19.4) | Reference |  | Reference |  |
| TST positive (N=288) | 104 (36.1) | 31 (31.3) | 73 (38.6) | 0.81 (0.57-1.15) | 0.230 |  |  |
| QFT Positive (N=222) | 36 (12.3) | 17 (16.8) | 19 (9.9) | 1.51 (1.01-2.27) | **0.046** | 1.54 (1.01-2.34) | **0.044** |
| TB contact | 117 (40.1) | 38 (37.6) | 79 (41.4) | 0.90 (0.65-1.25) | 0.539 |  |  |
| CXR suggestive of TB | 185 (63.4) | 71 (70.3) | 114 (59.7) | 1.37 (0.96-1.95) | **0.083** | 1.05 (0.63-1.75) | 0.860 |
| Mtb culture/ Xpert positive(N=276)^e^ | 18 (6.2) | 9 (8.9) | 9 (4.7) | 1.50 (0.92-2.46) | 0.108 |  |  |
| Urine LAM positive(N=166) | 17 (10.2) | 3 (5.7) | 14 (12.4) | 0.53 (0.18-1.51) | 0.232 |  |  |
| TBTx initiated^f^ | 127 (43.5) | 53 (52.5) | 74 (38.7) | 1.43 (1.05-1.97) | **0.025** | 1.22 (0.87 -1.72) | 0.257 |
| Positive response TBTx (N=127^g^ | 116 (91.3) | 51 (96.2) | 65 (87.8) | 2.42 (0.68-8.66) | 0.175 |  |  |
| **Abbreviations:** IQR: Interquartile range; WAZ ,weight for-age z score; WHZ, weight for-height z score; BMI, Body Mass Index (BMI) –for-age z score ; HUU, HIV unexposed uninfected; HEU, HIV exposed uninfected; CLHIV, children living with HIV; NIH, National Institutes of Health; TST, tuberculin skin test; QFT, QuantiFERON test; CXR, chest radiograph; Mtb, mycobacterium tuberculosis; Xpert, Xpert MTB/ULTRA; LAM, lipoarabinomannan; TBTx, TB treatment.  N: Number of participants with results; n: number of participants with positive results  ^a^ BMIz<-2 For the entire population  ^b^ Among children 5 years and under  ^c^ Among children 5 years and under: WHZ <-2 or MUAC <12·5 cm or WAZ<-2  ^d^ Persistent cough (>14 days), fever (>7 days), failure to thrive, or lethargy (>7 days)· Failure to thrive=wasted (WHZ<-2 or MUAC<12·5) or underweight (WHZ<-2) or Underweight (BMI <-2) at enrollment (growth trajectories unavailable before enrollment). ^e^ Sputum or gastric aspirate.  ^f^ Received TB treatment at enrollment or 2-weeks after enrollment  ^g^ Positive response to TB treatment after 2 weeks of enrollment  *Relative risk (RR) estimated using a generalized linear model (GLM) with log link and Poisson family  ^Ϊ^Adjusted for CXR in multivariate regression, including variables with p<0.1 from univariate analysis | | | | | | | |
