## Supplemental Figure 2: ROC for CRP to diagnose TB for "C-reactive protein for pulmonary tuberculosis screening and treatment response monitoring in children"

**Supplemental Figure 2: Receiver operating characteristic (ROC) for C-reactive protein to diagnose TB.**

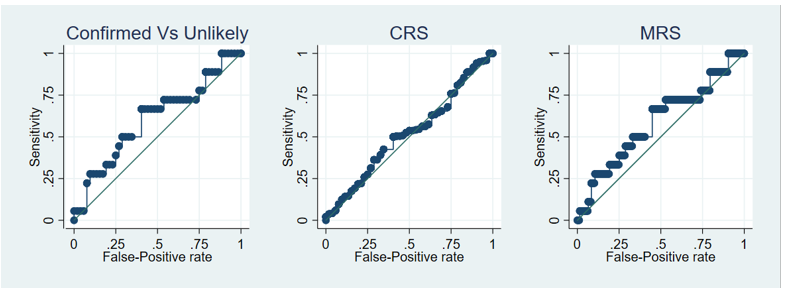

Reference Standards: Confirmed TB versus Unlikely TB, Composite reference standard (CRS)= Confirmed + Unconfirmed vs. Unlikely TB, Microbiological reference standard (MRS)= Confirmed vs. Unconfirmed + Unlikely
