## Supplemental Table 4: AUC under the ROC curve for CRP levels for "C-reactive protein for pulmonary tuberculosis screening and treatment response monitoring in children"

**Supplemental Table 4: Area under the ROC Curve for C-Reactive Protein Levels to**

**Detect Childhood TB**

|  | **AUC**  **(95% CI)** | **Cut off mg/L^a^** | **Sensitivity % (95%CI)** | **Specificity % (95%CI)** | **PPV** | **NPV** |
| --- | --- | --- | --- | --- | --- | --- |
| **Confirmed vs. Unlikely** | 0.61  (0.45, 0.77) | 2.28 | 66.7  (41.0, 86.7) | 59.6  (45.1, 73.0) | 36.4  (20.4, 54.9) | 83.8  (68.0, 93.8) |
| **MRS** | 0.59  (0.45, 0.74) | 2.44 | 66.7  (41.0, 86.7) | 55.1  (49.0, 61.1) | 8.9  (4.7, 15.0) | 96.2  (91.0, 98.6) |
| **CRS** | 0.51  (0.43, 0.60) | 2.10 | 50.0  (43.5, 56.5) | 59.6  (45.1, 73.0) | 85.1  (78.1, 90.5) | 20.5  (14.4, 27.9) |

**Abbreviations :** Receiver operating characteristic **(**ROC); Area under the ROC (AUC) ; Positive predictive value (PPV) ; Negative predictive value (NPV) ; Composite reference standard (CRS)= Confirmed + Unconfirmed vs. Unlikely TB, Microbiological reference standard (MRS)= Confirmed vs. Unconfirmed + Unlikely

; Composite reference standard on treatment (CRS on RX ) = Confirmed (on treatment) + Unconfirmed (on treatment) vs. Unlikely (not on treatment)

^a^ Optimal cut off for each of the reference standards
